# Causal effect of atrial fibrillation on brain white or grey matter volume: A Mendelian randomization study

**DOI:** 10.1101/2020.12.17.20248314

**Authors:** Sehoon Park, Soojin Lee, Yaerim Kim, Semin Cho, Kwangsoo Kim, Yong Chul Kim, Seung Seok Han, Hajeong Lee, Jung Pyo Lee, Soryoung Lee, Eue-Keun Choi, Kwon Wook Joo, Chun Soo Lim, Yon Su Kim, Dong Ki Kim

## Abstract

**Background:** Atrial fibrillation (AF) and brain volume loss are prevalent in older individuals. Further study investigating the causal effect of AF on brain volume is warranted.

**Methods:** This study was a Mendelian randomization (MR) analysis. The genetic instrument for AF was constructed from a previous genome-wide association study (GWAS) meta-analysis and included 537,409 individuals of European ancestry. The outcome summary statistics for quantile-normalized white or grey matter volume measured by magnetic resonance imaging were provided by the previous GWAS of 8426 white British UK Biobank participants. The main MR method was the inverse variance weighted method, supported by sensitivity MR analysis including MR-Egger regression and the weighted median method. The causal estimates from AF to white or grey matter volume were further adjusted for effects of any stroke or ischemic stroke by multivariable MR analysis.

**Results:** A higher genetic predisposition for AF (one standard deviation increase) was significantly associated with lower white matter volume [beta −0.128 (−0.208, −0.048)] but not grey matter volume [beta −0.041 (−0.101, 0.018)], supported by all utilized sensitivity MR analyses. The multivariable MR analysis indicated that AF is causally linked to lower white matter volume independent of the stroke effect.

**Conclusions:** AF is a causative factor for white matter volume loss. The effect of AF on grey matter volume was inapparent in this study. A future trial is necessary to confirm whether appropriate AF management can be helpful in preventing cerebral white matter volume loss or related brain disorders in AF patients.

## Introduction

Atrial fibrillation (AF) is the most common cardiac arrhythmia associated with risk of stroke, heart failure, dementia, and mortality^1^ and further contributes to a substantial socioeconomic burden.^2^ The prevalence of AF is substantially increasing along with the global aging trend.^3,4^

As AF is highly prevalent in elderly individuals, cognitive dysfunction or functional brain disorders, which are also common in elderly people, have been associated with AF.^5^ In addition, brain volume loss is related to persistent AF, along with low cerebral blood flow in AF patients.^6-8^ However, demonstration of the causal effect of AF on structural brain volume changes has yet to be performed. Because pathologic brain volume loss and AF are both common in older individuals with multiple comorbidities and diseases share risk factors, whether the observed low brain volume is a consequence of AF could hardly be answered by observational studies due to residual confounding effects. In addition, whether AF alone can cause brain volume loss even without stroke needs to be studied, as previous reports assessed the association between AF and dementia in stroke-free individuals, and brain volume loss was present before the identified first stroke event in AF patients.^5,9,10^ Such evidence for the causal effect of AF on brain volume and its mechanistic association with stroke would suggest that accelerated brain volume loss in AF patients may be ameliorated through appropriate AF management.

Mendelian randomization (MR) is an analytic method that can identify causal estimates with epidemiologic data.^11^ MR utilizes a genetic instrument that is fixed before birth; thus, the instrumented genetic predisposition is minimally affected by confounders or reverse causation. The significant association between genetic predisposition, which would result in a higher occurrence of the exposure of interest, and the outcome would suggest the presence of a causal effect of the exposure. MR has been widely introduced in the medical literature and has identified an important causal linkage between complex exposures and outcomes.

In this study, we performed a summary-level MR analysis to demonstrate the causal effects of AF on brain volume. By utilizing a recent large-scale genome-wide association study (GWAS) for white or grey matter brain volume, we derived causal estimates of AF on the two phenotypes. We also performed a multivariable MR analysis by adjusting for the effects of ischemic stroke to determine whether the effect of AF on brain volume is independent of the effect of stroke. We hypothesized that AF would decrease a certain type of brain volume with its distinct effect independent of stroke.

## Methods

### Ethical considerations

The study was approved by the institutional review boards of Seoul National University Hospital (No. E-2012-004-1177) and the UK Biobank consortium (application No. 53799). The study was performed in accordance with the Declaration of Helsinki. The study was performed by utilizing available public databases or GWAS summary statistics from previously published studies; thus, the requirement for informed consent was waived.

### Study setting

The study was a summary-level MR analysis based on published GWAS results (Figure 1). The causal estimates from AF to white or grey matter volume were initially tested. Next, additional GWAS results for any stroke or any ischemic stroke were also utilized, and the results were implemented for multivariable MR analysis adjusting for the effect of stroke on the causal estimates of AF on brain white or grey matter volume.

**Figure 1.**
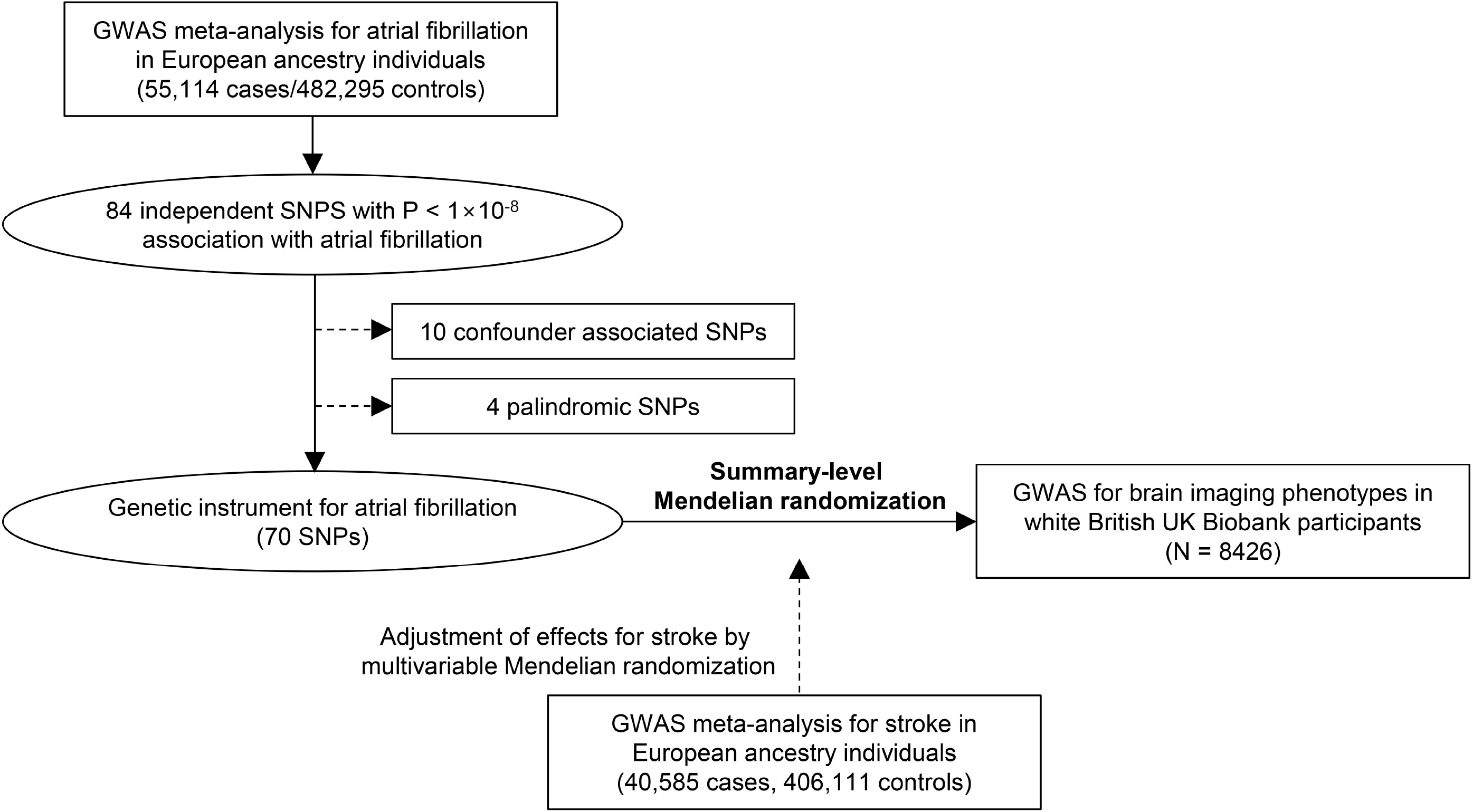
Study flow diagram.

### Genetic instrument for AF and MR assumptions

Data from a previous GWAS meta-analysis for AF were implemented for this study.^12^ As the outcome summary statistics for brain volume were limited to those with white British ancestry, we downloaded the summary statistics for AF in individuals of European ancestry, including 55,114 paroxysmal/persistent AF cases and 482,295 controls (URL: https://www.broadcvdi.org/). The study reported 84 independent sentinel SNPs not in linkage disequilibrium with P < 1×10^−8^ association strength with the AF phenotype. We utilized the summary statistics for the reported SNPs to construct the genetic instrument for AF.

MR analysis requires three key assumptions to be attained by the genetic instrument to demonstrate causal effects.^11^ First, the relevance assumption is that the genetic instrument should be strongly associated with the exposure of interest. The previous GWAS meta-analysis already provided SNPs with strong association strength with AF. We additionally tested the association strength in the UK Biobank data in 337138 unrelated individuals of white British ancestry who passed the sample quality control filter by calculating the degree of variance explained by polygenic score analysis with McFadden’s pseudo R-squared method. The polygenic score was calculated by multiplying the gene dosage matrix with the effect sizes of the genetic instrument by using PLINK 2.0 (version alpha 2.3).^13^ The AF outcome was collected from the hospital admission records or main causes of death, identified by an International Classification of Diseases (ICD)-10 diagnostic code of I48 or ICD-9 diagnostic code of 4273.^14^ Second, the independence assumption is that the genetic instrument should not be associated with confounders. To attain this assumption, we filtered out the SNPs with strong (P < 1×10^−5^) associations with hypertension, obesity, diabetes mellitus, dyslipidemia, and thyroid disorder in the abovementioned 337138 white British UK Biobank participants by performing a GWAS adjusted for age, sex, age×sex, age^2^, and the first 10 principal components. Further, in summary-level MR, we tested the presence of directional pleiotropy by identifying MR-Egger intercepts.^15^ In addition, we performed MR-Egger regression, which can yield valid causal estimates allowing 100% directional pleiotropy, and the weighted median method, which allows up to 50% invalid instrumented weights.^16^ Third, the exclusion restriction assumption is that the causal effect of interest should be through the studied exposure. Although a formal test for this assumption is not yet possible, the utilized median-based method can relax this assumption for up to 50% of the instrumented weights, providing sensitivity analysis for the attainment of this assumption.^16^

### Outcome GWAS for brain volume

A previous GWAS for brain imaging traits in the UK Biobank was performed in 8428 subjects of white British ancestry.^17^ The study identified that brain imaging phenotypes are mostly genetically trackable and reported functionally relevant genetic information associated with brain structures. The brain volume was measured by magnetic resonance imaging, and the study provided summary statistics for quantile-normalized brain volume phenotypes. The study also reported results for additionally normalized brain volumes corrected for various potential confounders, including age, sex, image device-associated factors, head motion, head position, and head size. We downloaded the summary statistics for normalized and unnormalized white or grey matter volume for the outcome data in the summary-level MR. The study details are available in a previously published article.^17^

### Summary-level MR analysis methods

In the summary-level MR, the variants that were palindromic were excluded during the harmonization of the summary statistics.^18^ The main MR method was the fixed-effects inverse variance weighted method, and additional pleiotropy-robust sensitivity MR analyses were performed, as the inverse variance weighted method requires the absence of pleiotropy. First, MR-Egger regression, which allows 100% pleiotropy and still yields valid causal estimates, was performed with the test for the presence of directional pleiotropy or MR-Egger intercept. Second, the weighted median method was implemented, which derives valid causal estimates even under conditions when up to 50% invalid instrumented weights are present.^16^ Finally, MR-PRESSO, which detects and corrects the effects of outliers, was performed when the test for global heterogeneity was significant.^19^ The summary-level MR analysis was performed by the TwoSampleMR package in R (version 4.0.2, the R foundation).^20^

### Sensitivity MR analysis in concerns of the presence of overlapping samples

As the GWAS for AF included UK Biobank participants, the study population in the GWAS for brain volume overlapped. Such overlapping samples in summary-level MR may potentially cause a bias towards observational findings, particularly when the genetic instrument has weak power.^21^ Following the previous literature investigating this issue,^21^ we performed another sensitivity analysis with modification of the genetic instrument, including fewer SNPs with a stronger association with the exposure of interest. In the analysis, we filtered SNPs within a range of thresholds for the association strength with AF (P < 1×10^−9^ to < 1×10^−18^), repeatedly performed summary-level MR analysis by the inverse variance weighted method, and performed the MR-Egger test to detect the presence of directional pleiotropy.

### MR analysis with stroke

Multivariable MR analysis is possible with direct adjustment for the effects of other phenotypes of the utilized genetic instrument.^22^ Considering that AF is well known to be associated with stroke, we performed additional multivariable MR analysis adjusting for the effects of stroke in the causal estimates of AF on brain volume. We hypothesized that the effects of AF on brain volume would be present independent of the effects of stroke. The summary statistics of the GWAS meta-analysis for any stroke or any ischemic stroke phenotype were available in a previous study.^23^ We downloaded the summary statistics of 446,696 individuals of European ancestry, including 40,585 stroke cases (https://www.megastroke.org/). The multivariable MR analysis derived the causal estimates of AF on white or grey matter brain volume adjusted for the effect of any stroke or any ischemic stroke. The analysis was performed by the MVMR package in R.^24^

In addition, as a supplemental analysis, we performed a summary-level MR with the 32 SNPs reported in the previous GWAS meta-analysis associated with any type of stroke as the genetic instrument for stroke. Although some SNPs were identified in the trans-ancestry individuals and the related stroke subtypes were different, all SNPs were utilized, as the number of genome-wide significant SNPs was small. We performed a summary-level MR analysis with the 32 SNPs and a sensitivity MR with the genetic instrument disregarding possible confounder-associated SNPs as in the abovementioned methods.

## Results

### Genetic instrument for AF

Among the 84 SNPs reported in the previous GWAS meta-analysis of individuals of European ancestry for their strong association with AF, 4 SNPs were disregarded due to association with possible confounders, and 10 SNPs were filtered out because they were palindromic, resulting in the use of 70 SNPs to construct the genetic instrument for AF (Supplemental Table 1). Among the 337138 white British ancestry individuals of the UK Biobank, the median age was 58 years old, and 46.3% were males. From this group, we identified 15,446 (4.6%) AF cases. The polygenic score calculated by the genetic instrument was strongly associated (P < 2×10^−16^) with phenotypical AF, explaining 2.4% of the variance in the trait.

### Causal estimates from AF to brain volume

The summary statistics of the utilized genetic instruments for the outcome traits are presented in Supplemental Table 2. The summary-level MR results indicated that genetic predisposition for AF was significantly associated with lower white matter volume, both normalized and unnormalized (Figure 2, Figure 3, and Table 1). The results were also significant in the MR-Egger regression analysis and the weighted median methods. The MR-PRESSO test for global heterogeneity did not identify correctable outlier effects; thus, the MR-PRESSO analysis was unnecessary. The MR-Egger intercepts were nonsignificant, indicating the absence of a significant directional pleiotropic effect in the results. On the other hand, the causal estimates of grey matter volume did not identify a significant association between genetic predisposition for AF and normalized or unnormalized volume by the MR analysis utilized.

**Table 1.** Causal estimates from atrial fibrillation on brain or white matter volume of the UK Biobank participants by summary-level Mendelian randomization.

| Outcome | MR method | MR-Egger intercept P value | beta (95% CI) | P value |
| --- | --- | --- | --- | --- |
| White matter volume (normalized) | Inverse variance weighted | 0.102 | -0.059 (-0.111, -0.007) | 0.025 |
|  | MR-Egger |  | -0.204 (-0.386, -0.022) | 0.031 |
|  | Weighted median |  | -0.097 (-0.175, -0.018) | 0.016 |
| White matter volume (unnormalized) | Inverse variance weighted | 0.090 | -0.030 (-0.057, -0.003) | 0.028 |
|  | MR-Egger |  | -0.107 (-0.200, -0.014) | 0.028 |
|  | Weighted median |  | -0.048 (-0.09, -0.006) | 0.025 |
| Grey matter volume (normalized) | Inverse variance weighted | 0.122 | -0.014 (-0.053, 0.024) | 0.460 |
|  | MR-Egger |  | -0.112 (-0.243, 0.018) | 0.095 |
|  | Weighted median |  | -0.011 (-0.065, 0.044) | 0.695 |
| Grey matter volume (unnormalized) | Inverse variance weighted | 0.100 | -0.009 (-0.035, 0.018) | 0.522 |
|  | MR-Egger |  | -0.078 (-0.165, 0.008) | 0.081 |
|  | Weighted median |  | -0.009 (-0.048, 0.030) | 0.658 |

MR = Mendelian randomization, CI = confidence interval
The effect sizes of the causal estimates were from one standard deviation increase of the genetic predisposition for atrial fibrillation to the quantile normalized brain volume phenotype (a one-standard deviation unit).
MR-PRESSO analysis was performed but as MR-PRESSO global test for heterogeneity did not identify correctable effects from outliers, the causal estimates were same as the inverse variance weighted method.

**Figure 2.**
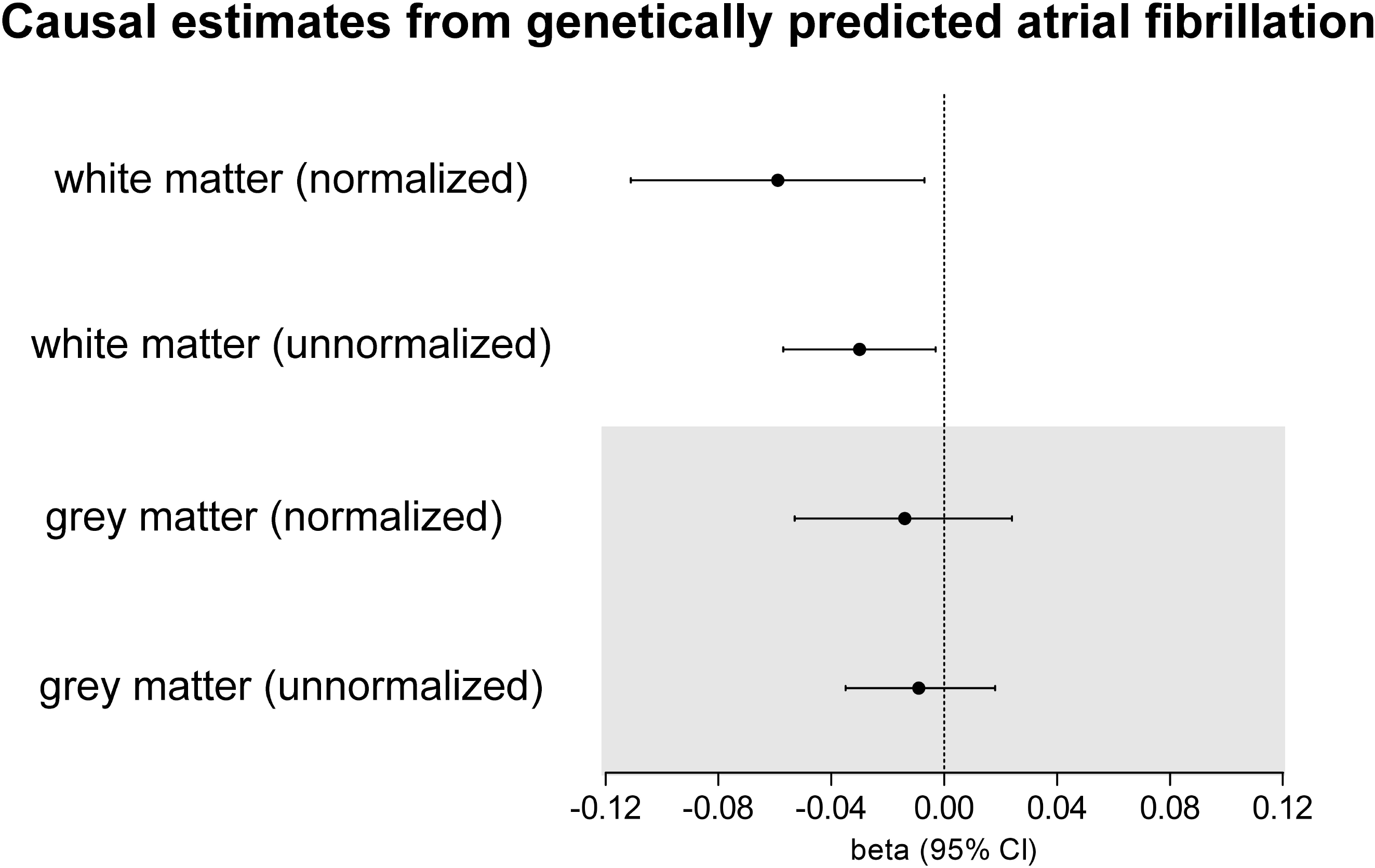
Causal estimates from atrial fibrillation on brain white or grey matter volume. The causal estimates were derived by inverse variance weighted method. The genetic instrument of 70 single nucleotide polymorphisms predicted atrial fibrillation.

**Figure 3.**
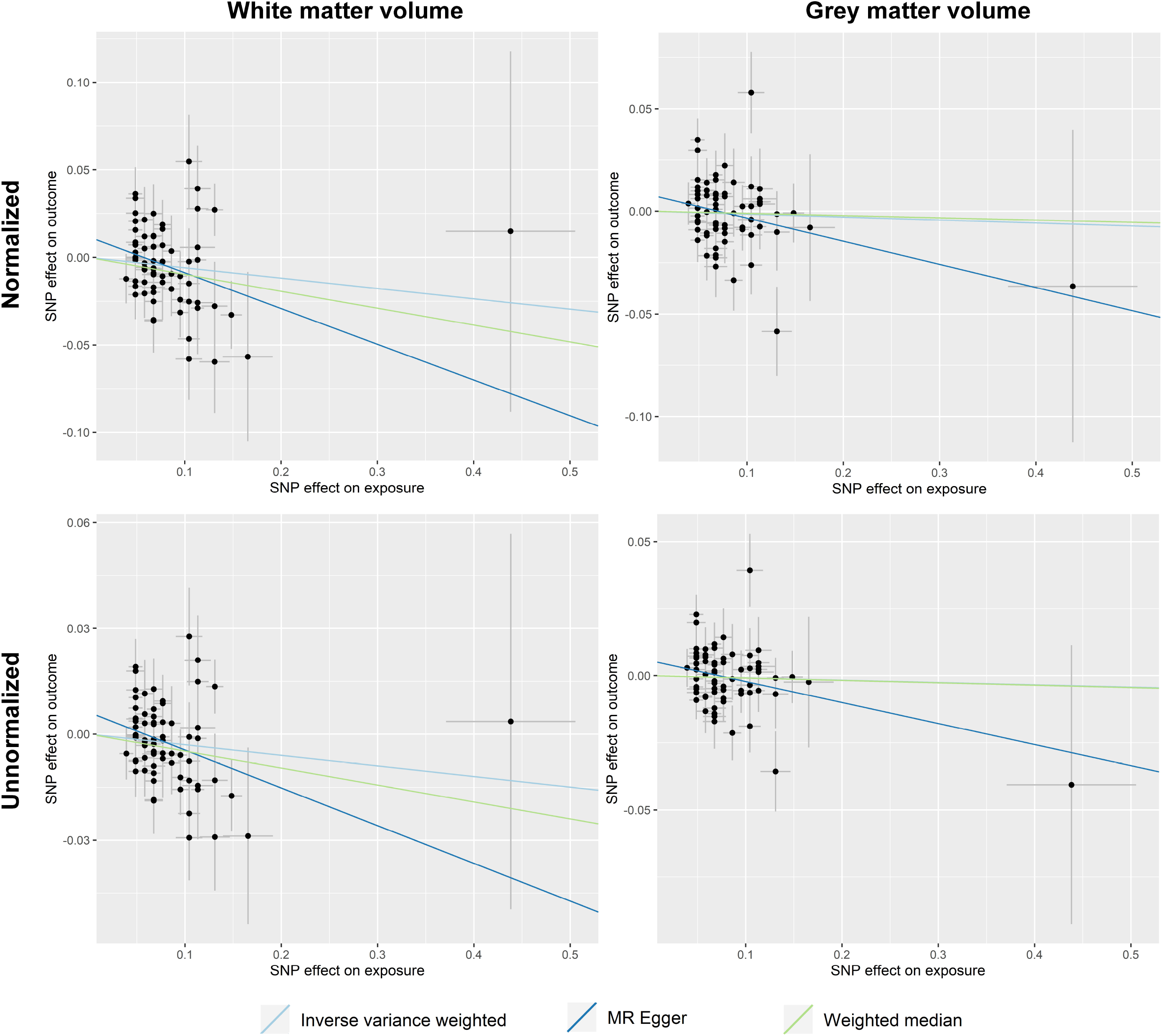
Scatter plot of the Mendelian randomization analysis showing the causal estimates from atrial fibrillation on brain volume phenotypes.

### Sensitivity analysis results with fewer but stronger SNPs

When we applied a more stringent association strength filter for the SNPs, even after reducing ∼80% of SNPs by applying the threshold P < 1×10^−18^, the genetic predisposition for AF was significantly associated with low white matter volume (Supplemental Table 3). However, the causal estimates of AF on grey matter volume were mostly nonsignificant in this sensitivity analysis.

### MR results with stroke

One of the SNPs, rs187585530 with a regression beta effect size of over 0.4 for AF and an effect allele frequency of 0.005, was unavailable in the summary statistics for stroke and thus disregarded in the multivariable MR analysis. The summary statistics of the 69 SNPs utilized as the genetic instrument for AF and their association with any stroke or ischemic stroke are presented in Supplemental Table 4. When we adjusted for the effects of any stroke or ischemic stroke, the causal estimates of AF on lower white matter volume remained significant (Table 2). The results were similar for both normalized and unnormalized white matter volumes. Again, the causal estimates in the multivariable MR analysis were nonsignificant for grey matter volume.

**Table 2.** Multivariable Mendelian randomization analysis showing the causal estimates from atrial fibrillation on brain volume phenotypes adjusted for effects from stroke.

| Outcome | Phenotype effect adjusted | beta (95% CI) | P |
| --- | --- | --- | --- |
| White matter volume (normalized) | Any stroke | -0.128 (-0.208, -0.048) | 0.029 |
|  | Any ischemic stroke | -0.136 (-0.215, -0.056) | 0.001 |
| White matter volume (unnormalized) | Any stroke | -0.059 (-0.101, -0.016) | 0.008 |
|  | Any ischemic stroke | -0.062 (-0.104, -0.02) | 0.005 |
| Grey matter volume (normalized) | Any stroke | -0.041 (-0.101, 0.018) | 0.179 |
|  | Any ischemic stroke | -0.030 (-0.090, 0.030) | 0.324 |
| Grey matter volume (unnormalized) | Any stroke | -0.030 (-0.070, 0.009) | 0.136 |
|  | Any ischemic stroke | -0.022 (-0.062, 0.018) | 0.276 |

CI = confidence interval
Multivariable Mendelian randomization analysis was performed by inverse variance weighted method adjusted for the effects for stroke phenotypes of the utilized 69 SNPs for the genetic instrument of atrial fibrillation.
The effect sizes of the causal estimates were from one standard deviation increase of the genetic predisposition for atrial fibrillation to the quantile normalized brain volume phenotype (a one-standard deviation unit).

The summary statistics of the genetic instrument for the stroke trait are presented in Supplemental Table 5. Among them, 2 SNPs were palindromic, and 3 SNPs did not overlap with the outcome summary statistics; thus, stroke was predicted by 27 SNPs. When the causal estimates from the genetic instrument for SNPs on brain volume were tested, the causal estimates were nonsignificant for both white and grey matter volumes (Supplemental Table 6). Additional exclusion of the 9 SNPs associated with possible confounders again yielded nonsignificant results.

## Discussion

This study identified that genetic predisposition for AF is significantly associated with lower white matter volume but not grey matter volume. With our efforts to attain the MR assumptions, our study supports that AF is a causative factor for decreased white matter volume. The suggested effects were independent of the effect of stroke in the multivariable MR analysis. The causal effect of stroke on brain volume was inapparent in the current results.

An observational association between AF and functional brain disorders, representatively dementia, has been reported.^5,9^ A recent study identified that baseline AF was associated with an incident risk of dementia, which was also identified in patients without a history of stroke.^9^ A previous study reported that AF was associated with lower brain volume in nondemented elderly individuals.^7^ The association between AF and low brain volume was significant regardless of the presence of cerebral infarcts,^8^ suggesting that low brain blood flow related to AF or other common risk factors may have driven brain volume loss in patients with AF.^10^ However, as AF and structural or functional brain disorders are prevalent in elderly individuals with many comorbidities, such observational findings are prone to residual confounding effects or reverse causation. Moreover, as brain volume is a relatively unique phenotype available in a large number of individuals and rarely sequentially measured, a study investigating the causal effect of AF on brain volume is very difficult to perform with a conventional observational design. In this study, we implemented MR analysis to test causal estimates from exposure and predicted complex outcomes using the genetic instrument. Finally, we identified that AF causally decreased white matter volume. The identified causal estimates were independent of the effect from ischemic or any stroke; thus, the study suggests that AF causes brain white matter volume loss independent of stroke.

AF causes hemodynamic compromises in the brain.^8^ Moreover, AF causes atrial dysfunction with impaired ventricular filling, consequently adversely affecting cardiac output.^25^ AF is also closely associated with adverse neurohormonal responses, including activation of the renin-angiotensin-aldosterone system,^26,27^ which mechanistically causes cerebral vasoconstriction.^28^ Brain volume loss in AF patients showed greater accentuation in those with persistent AF than in those with paroxysmal AF, suggesting that the cumulative burden of AF might influence decreased brain volume.^7^ Further, a previous study suggested that white matter is more prone to ischemic injury,^29^ supporting our findings that genetically predicted AF was significantly associated with low white matter volume but not with grey matter volume. That AF is associated with higher odds of silent cerebral infarction supports that long-term subclinical brain hemodynamic compromise due to AF may be the cause of white matter volume loss.^30^ However, the confirmation of the mechanism of the identified causal effects from AF on brain white matter volume is beyond the possibility of this MR analysis, and a future study may investigate the hemodynamic effect of AF on brain blood flow and related consequences on cerebral volume change.

Based on the study results, as AF is a causative factor for brain white matter volume loss, appropriate AF management might delay white matter volume loss. Considering that causal estimates were independent of ischemic stroke effects, appropriate anticoagulation therapy alone may not be sufficient to delay brain volume loss. A future study would be needed to test our hypothesis that early rhythm-control therapy in patients with AF would decrease the risk of brain volume loss in patients with AF. Early rhythm-control therapy recently showed a decreased risk of cardiovascular outcomes compared to usual care in patients with early atrial fibrillation and cardiovascular conditions.^31^ In addition, monitoring brain volume would be important in high-risk patients, as white matter abnormalities are common in the early stages of dementia.^32-34^ Considering that subclinical AF and early brain disorders are often underdiagnosed, AF screening in the high-risk elderly population with brain imaging would be considered for future study to facilitate early diagnosis of important comorbidities.

There are several limitations to this study. First, the UK Biobank cohort overlapped in the populations for genetic instrument development and outcome measurement, which has been reported to potentially cause a bias towards observational findings.^21^ Although we performed sensitivity analyses to demonstrate that the findings are not from type 1 error, such overlapping sample issues should be noted in summary-level GWAS based on data from large genetic consortia. Second, the nonsignificant causal effects from a stroke on brain volume are not confirmative. The instrumented SNPs to genetically predict stroke were introduced from a transethnic population due to data availability, and the possibility of weak instrument bias remains, which is towards the null in summary-level MR. Moreover, even though a direct effect on structural brain volume was unidentified, stroke is an undebatable cause of functional and structural brain disorders. Thus, the finding should be interpreted to be supportive for the causative effect of AF on brain volume was strong even when considering the effect of stroke but should not disregard the importance of stroke in pathologic changes in the brain. Third, the study is mainly based on data from individuals of European ancestry; thus, generalizability to other ethnic populations is not secured. Fourth, although the previous GWAS meta-analysis provided genetic variants strongly associated with AF, the AF phenotype included both paroxysmal and persistent AF. Thus, whether the effects of AF are different according to the subtypes could not be studied herein. Last, a quantitative interpretation of the results is limited because the degree of the clinical effect of AF on brain white matter volume may be different from the genetic findings in this study.^35^

In conclusion, AF causally decreases brain white matter volume, independent of the effect of ischemic stroke. A future study is warranted to investigate whether appropriate AF management may result in delayed progression of white matter volume loss in AF patients.

## Data Availability

Data for this study will be made available through the UK Biobank. The implemented GWAS summary statistics are available in the public domain.

## Acknowledgements

The study was based on the data provided by the UK Biobank consortium (application No. 53799), the Cardiovascular Disease Knowledge Portal, and the MEGASTROKE project. The MEGASTROKE project received funding from sources specified at http://www.megastroke.org/acknowledgments.html. We thank the investigators of the previous studies who provided the valuable genetic summary statistics for this study.

## Funding

This work was supported by the Industrial Strategic Technology Development Program - Development of bio-core technology (10077474, Development of early diagnosis technology for acute/chronic renal failure) funded by the Ministry of Trade, Industry & Energy (MOTIE, Korea) and a grant from SNU R&DB Foundation (800-20190571). The study was performed independently by the authors.

## Conflicts of interest

EKC: research grants from Bayer, BMS/Pfizer, Biosense Webster, Chong Kun Dang, Daiichi-Sankyo, Dreamtech Co. Ltd., Medtronic, Samjinpharm, Sanofi-Aventis, Seers Technology, Skylabs, and Yuhan. No fees are received personally.

