## Supplemental Table 3 and 6 for "Causal effect of atrial fibrillation on brain white or grey matter volume: A Mendelian randomization study"

**Supplemental Table 3.** Sensitivity analysis applying ranges of thresholds to filter in SNPs in the genetic instrument.

| Outcome | P value threshold to filter SNPs | N of SNPS remained | beta (95% CI) | P by inverse variance weighted | MR-Egger intercept P |
|---|---|---|---|---|---|
| White matter volume (normalized) | $< 1\times10^{-9}$ | 57 | -0.064 (-0.119, -0.01) | 0.02 | 0.095 |
| | $< 1\times10^{-10}$ | 47 | -0.092 (-0.149, -0.035) | 0.002 | 0.992 |
| | $< 1\times10^{-11}$ | 40 | -0.087 (-0.147, -0.028) | 0.004 | 0.907 |
| | $< 1\times10^{-12}$ | 38 | -0.088 (-0.148, -0.028) | 0.004 | 0.922 |
| | $< 1\times10^{-13}$ | 32 | -0.091 (-0.154, -0.028) | 0.005 | 0.668 |
| | $< 1\times10^{-14}$ | 29 | -0.091 (-0.156, -0.026) | 0.006 | 0.631 |
| | $< 1\times10^{-15}$ | 28 | -0.09 (-0.156, -0.024) | 0.007 | 0.615 |
| | $< 1\times10^{-16}$ | 24 | -0.074 (-0.143, -0.005) | 0.034 | 0.444 |
| | $< 1\times10^{-17}$ | 19 | -0.075 (-0.149, -0.002) | 0.045 | 0.382 |
| | $< 1\times10^{-18}$ | 18 | -0.085 (-0.16, -0.011) | 0.025 | 0.560 |
| White matter volume (unnormalized) | $< 1\times10^{-9}$ | 57 | -0.033 (-0.061, -0.005) | 0.022 | 0.091 |
| | $< 1\times10^{-10}$ | 47 | -0.047 (-0.076, -0.017) | 0.002 | 0.990 |
| | $< 1\times10^{-11}$ | 40 | -0.045 (-0.075, -0.014) | 0.004 | 0.922 |
| | $< 1\times10^{-12}$ | 38 | -0.044 (-0.076, -0.013) | 0.005 | 0.920 |
| | $< 1\times10^{-13}$ | 32 | -0.046 (-0.078, -0.013) | 0.006 | 0.684 |
| | $< 1\times10^{-14}$ | 29 | -0.046 (-0.079, -0.012) | 0.008 | 0.632 |
| | $< 1\times10^{-15}$ | 28 | -0.045 (-0.079, -0.011) | 0.009 | 0.616 |
| | $< 1\times10^{-16}$ | 24 | -0.037 (-0.072, -0.001) | 0.042 | 0.431 |
| | $< 1\times10^{-17}$ | 19 | -0.038 (-0.076, 0.000) | 0.053 | 0.337 |
| | $< 1\times10^{-18}$ | 18 | -0.043 (-0.082, -0.004) | 0.030 | 0.507 |
| Grey matter volume (normalized) | $< 1\times10^{-9}$ | 57 | -0.016 (-0.056, 0.024) | 0.438 | 0.151 |
| | $< 1\times10^{-10}$ | 47 | -0.030 (-0.072, 0.012) | 0.164 | 0.945 |
| | $< 1\times10^{-11}$ | 40 | -0.032 (-0.076, 0.012) | 0.150 | 0.939 |
| | $< 1\times10^{-12}$ | 38 | -0.029 (-0.074, 0.015) | 0.196 | 0.873 |
| | $< 1\times10^{-13}$ | 32 | -0.045 (-0.092, 0.002) | 0.058 | 0.847 |
| | $< 1\times10^{-14}$ | 29 | -0.048 (-0.096, 0.000) | 0.049 | 0.990 |
| | $< 1\times10^{-15}$ | 28 | -0.046 (-0.095, 0.002) | 0.062 | 0.942 |
| | $< 1\times10^{-16}$ | 24 | -0.044 (-0.094, 0.007) | 0.091 | 0.903 |
| | $< 1\times10^{-17}$ | 19 | -0.04 (-0.094, 0.014) | 0.149 | 0.428 |
| | $< 1\times10^{-18}$ | 18 | -0.037 (-0.093, 0.018) | 0.184 | 0.491 |
| Grey matter volume (unnormalized) | $< 1\times10^{-9}$ | 57 | -0.009 (-0.037, 0.018) | 0.504 | 0.107 |
| | $< 1\times10^{-10}$ | 47 | -0.019 (-0.048, 0.010) | 0.202 | 0.969 |
| | $< 1\times10^{-11}$ | 40 | -0.020 (-0.050, 0.010) | 0.188 | 0.955 |
| | $< 1\times10^{-12}$ | 38 | -0.018 (-0.048, 0.013) | 0.256 | 0.81 |
| | $< 1\times10^{-13}$ | 32 | -0.028 (-0.060, 0.004) | 0.085 | 0.793 |
| | $< 1\times10^{-14}$ | 29 | -0.031 (-0.064, 0.002) | 0.065 | 0.989 |
| | $< 1\times10^{-15}$ | 28 | -0.029 (-0.062, 0.004) | 0.085 | 0.924 |
| | $< 1\times10^{-16}$ | 24 | -0.028 (-0.062, 0.007) | 0.119 | 0.915 |
| | $< 1\times10^{-17}$ | 19 | -0.026 (-0.063, 0.011) | 0.168 | 0.496 |
| | $< 1\times10^{-18}$ | 18 | -0.025 (-0.062, 0.013) | 0.202 | 0.558 |

CI = confidence interval

The SNPs reaching the designated P value threshold remained in the genetic instrument for atrial fibrillation in each analysis. The causal estimates were derived by the inverse variance weighted method, and the MR-Egger intercept P values were calculated to detect the presence of significant pleiotropy.

**Supplemental Table 6.** Causal estimates from stroke on brain white or grey matter volume.

| Outcome | Genetic instrument | MR method | MR-Egger intercept P value | beta (95% CI) | P value |
|---|---|---|---|---|---|
| Genetic instrument for stroke including all genome-wide significant SNPs reported in the previous GWAS (N = 27) | White matter volume (normalized) | Inverse variance weighted | 0.931 | -0.003 (-0.059, 0.054) | 0.925 |
| | | MR-Egger | | -0.006 (-0.103, 0.091) | 0.903 |
| | | Weighted median | | 0.007 (-0.075, 0.089) | 0.864 |
| | White matter volume (unnormalized) | Inverse variance weighted | 0.845 | -0.002 (-0.031, 0.027) | 0.877 |
| | | MR-Egger | | -0.006 (-0.055, 0.043) | 0.807 |
| | | Weighted median | | 0.004 (-0.038, 0.045) | 0.865 |
| | Grey matter volume (normalized) | Inverse variance weighted | 0.114 | 0.019 (-0.022, 0.061) | 0.36 |
| | | MR-Egger | | -0.024 (-0.091, 0.043) | 0.482 |
| | | Weighted median | | 0.006 (-0.054, 0.066) | 0.841 |
| | Grey matter volume (unnormalized) | Inverse variance weighted | 0.076 | 0.013 (-0.016, 0.041) | 0.377 |
| | | MR-Egger | | -0.021 (-0.067, 0.025) | 0.375 |
| | | Weighted median | | -0.007 (-0.049, 0.035) | 0.742 |
| Genetic instrument for stroke after excluding confounder-associated SNPs (N = 18) | White matter volume (normalized) | Inverse variance weighted | 0.930 | 0.011 (-0.050, 0.073) | 0.719 |
| | | MR-Egger | | 0.015 (-0.085, 0.115) | 0.773 |
| | | Weighted median | | 0.014 (-0.066, 0.095) | 0.729 |
| | White matter volume (unnormalized) | Inverse variance weighted | 0.963 | 0.005 (-0.027, 0.037) | 0.749 |
| | | MR-Egger | | 0.004 (-0.047, 0.056) | 0.874 |
| | | Weighted median | | 0.007 (-0.039, 0.052) | 0.774 |
| | Grey matter volume (normalized) | Inverse variance weighted | 0.259 | 0.014 (-0.032, 0.06) | 0.547 |
| | | MR-Egger | | -0.02 (-0.094, 0.053) | 0.594 |
| | | Weighted median | | 0.006 (-0.059, 0.071) | 0.848 |
| | Grey matter volume (unnormalized) | Inverse variance weighted | 0.176 | 0.011 (-0.020, 0.042) | 0.492 |
| | | MR-Egger | | -0.018 (-0.068, 0.033) | 0.503 |
| | | Weighted median | | 0.000 (-0.043, 0.042) | 0.984 |

CI = confidence interval
